# Randomized, placebo-controlled, pilot clinical study evaluating acute Niagen®+ IV and NAD+ IV in healthy adults

**DOI:** 10.1101/2024.06.06.24308565

**Authors:** Jessie Hawkins, Rebecca Idoine, Jun Kwon, Andrew Shao, Elizabeth Dunne, Elizabeth Hawkins, Kayla Dawson, Yasmeen Nkrumah-Elie

## Abstract

**Background:** Nicotinamide riboside (NR) is a promising compound for augmenting the intracellular NAD+ pool, potentially mitigating age-related decline and associated conditions. While oral NR supplementation has demonstrated safety and bioavailability in multiple animal and human studies, the effects of intravenous NR (NR IV) are far less understood. Until now, pharmaceutical grade NR was not available for injection research.

**Objectives:** Given that intravenous administration may offer advantages in certain conditions and contexts, a systematic investigation of the clinical effects of NR IV is warranted.

**Methods:** The present randomized, double-blinded, placebo-controlled, pilot clinical study was initiated with the primary aim of investigating the safety, tolerability, and the blood NAD+-boosting efficacy of an acute, single dose of NR IV (500 mg, test), NAD+ IV (500 mg, active comparator), oral NR (500 mg, bridge), and saline IV (placebo control) in generally healthy adult participants. The study consisted of two parts; data from 37 and 16 participants in the first and second phases, respectively, were analyzed.

**Results:** No significant differences in vital signs were detected across groups. In comparison to NAD+ IV, NR IV was associated with fewer and less severe adverse experiences during the infusion; no attributable adverse events were reported through the 14-day follow-up period for any treatment groups. Further, the mean tolerable infusion time for NR IV was 75% less than that of NAD+ IV. No clinically meaningful changes in blood chemistry markers were described in the NR IV condition, whereas an increase in white blood cell counts and neutrophils was observed in the NAD+ IV condition, suggesting the presence of an inflammatory response. Finally, NR IV appeared to promote the most robust increases in NAD+ concentration as measured by dried blood spot analyses, with peak NAD+ levels increasing by 20.7% relative to baseline, and acutely outperforming NAD+ IV (p <0.01) and oral NR (p<0.01) at the 3-hr timepoint.

**Conclusion:** This is the first study to clinically evaluate NR IV. Overall, acute intravenous infusions of 500 mg NR were safe in the study participants with no attributable adverse events and only minor and transient infusion-related experiences. In comparison to NAD+ IV, NR IV had a faster infusion time with superior tolerability. At 3 hours post-infusion, blood NAD+ levels were significantly higher in the NR IV group compared to the NAD+ IV group. Future studies in larger populations are needed to validate these results.

## Introduction

Nicotinamide adenine dinucleotide (NAD+, **Figure 1A**) is an essential coenzyme present in all living cells. Due to its involvement in hundreds of reduction-oxidation (redox) reactions, NAD+ plays a fundamental role in metabolic pathways responsible for fulfilling bioenergetic needs.

**Figure 1.**
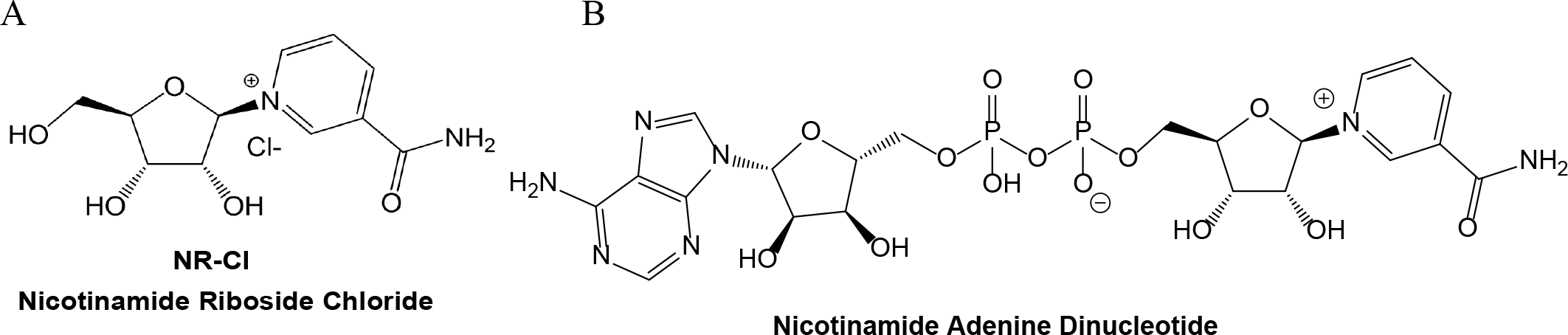
Chemical Structures of Nicotinamide Riboside Chloride and Nicotinamide Adenine Dinucleotide.

**Figure 2.**
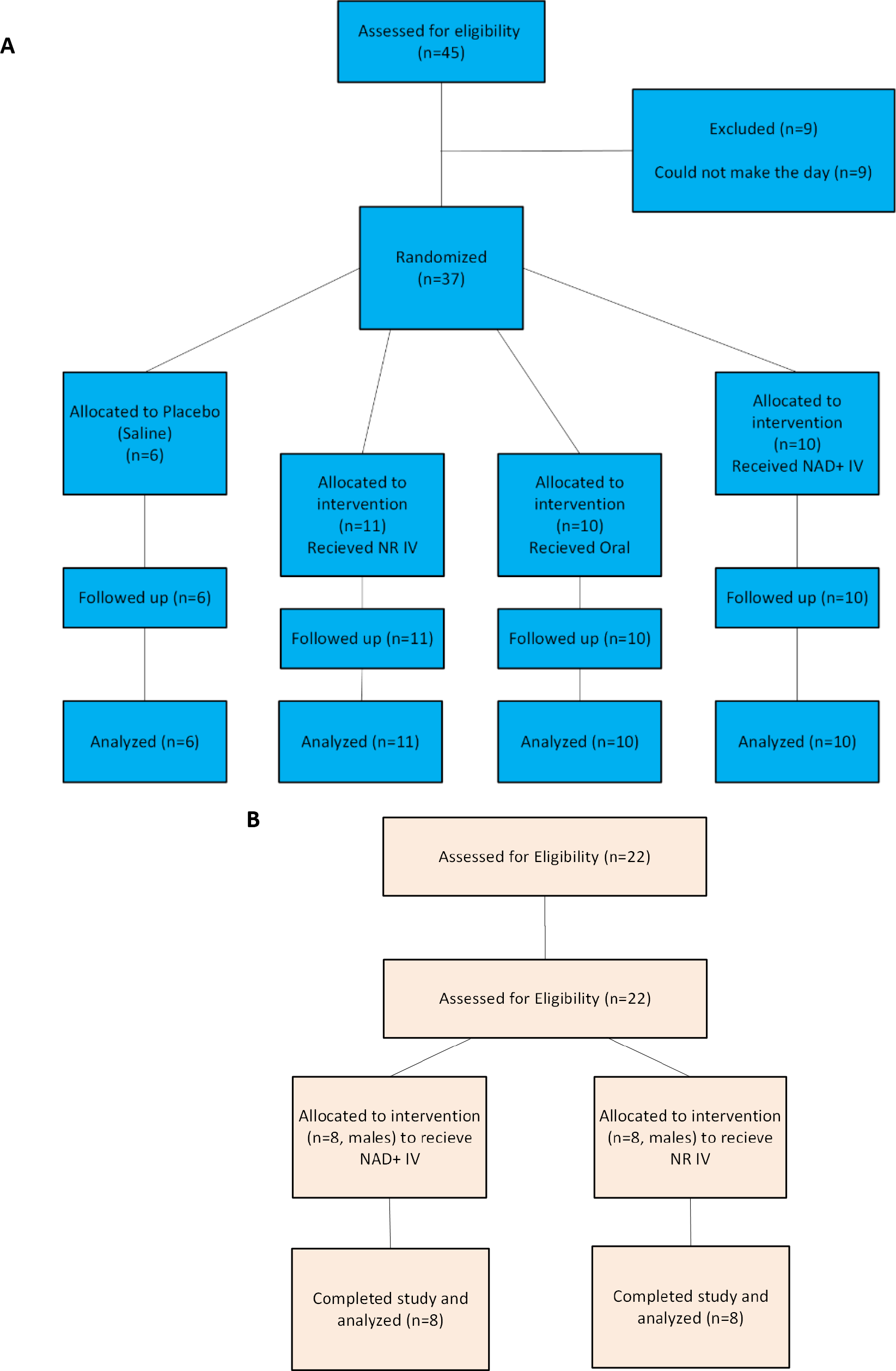
Study flow diagram for study 1 (A) and study 2 (B).

NAD+ functions as a substrate in non-redox enzymatic reactions, including, those controlling DNA repair, gene expression, and calcium signaling.

Aging is accompanied by reductions in cellular and tissue NAD+ concentrations, driven at least in part by an accelerated rate of CD38-mediated NAD+ degradation (Camacho-Pereira *et al*., 2016; McReynolds *et al*., 2021; Chini *et al*., 2024). The resultant decline in NAD+ levels has been casually implicated with the development of mitochondrial dysfunction, the biological hallmarks of aging, and may contribute to age-associated disorders and abnormalities, such as neurodegeneration, hypertension, and chronic inflammation (Fang *et al*., 2017; Covarrubias *et al*., 2021). NAD+ homeostasis is also disturbed in conditions of metabolic stress, including heart failure, diabesity, central and peripheral neurodegeneration, mitochondrial disease, alcoholic liver disease, postpartum, coronavirus infection (Samuel A.J. Trammell *et al*., 2016; Hamity *et al*., 2017; Vaur *et al*., 2017; Diguet *et al*., 2018; Liu *et al*., 2018; Ear *et al*., 2019; Parker *et al*., 2020; Pirinen *et al*., 2020; Heer *et al*., 2021). On the other hand, interventions that boost NAD+ availability have repeatedly been demonstrated to rescue defects associated with the loss of NAD+ homeostasis and improve physiological function (Cantó *et al*., 2012; Zhang *et al*., 2016; Martens *et al*., 2018; Elhassan *et al*., 2019).

While NAD+ is commercially available as dietary supplement and intravenous drug products, as a pyridine nucleotide, NAD+ itself is unable to undergo direct intestinal absorption or cellular uptake intact upon exogenous administration (Nikiforov *et al*., 2011). The majority of NAD+ is instead hydrolyzed in the extracellular environment to nicotinamide mononucleotide (NMN), which in turn is further cleaved by CD73 to form nicotinamide riboside (NR), a portion of which may be further degraded to nicotinamide and nicotinic acid (Nikiforov *et al*., 2011; Anthony A. Sauve *et al*., 2023). NR (**Figure 1B**) is readily taken up by cells via equilibrative nucleoside transporters and directed toward NAD+ biosynthesis in a two-step process involving the nicotinamide riboside kinase enzymes (Bieganowski and Brenner, 2004; Nikiforov *et al*., 2011; Ratajczak *et al*., 2016; Kropotov *et al*., 2021). Therefore, the provision of exogenous NR, rather than NAD+ itself, appears to be more efficient for augmenting intracellular NAD+ concentrations. Indeed, oral NR administration has been described to be safe and effective in raising NAD+ and has shown promise against neurodegenerative conditions and other age-related disorders (Samuel A. J. Trammell *et al*., 2016; Elhassan *et al*., 2019; Brakedal *et al*., 2022; Berven *et al*., 2023; Biţă *et al*., 2023) In spite of the aforementioned limitations concerning the provision of exogenous NAD+, the intravenous (IV) administration of NAD+ (‘NAD+ IV’) has gained popularity in recent years and is available in thousands of boutique medical and hydration clinics globally. Initially described in the clinical literature in 1961 for use in the treatment of multiple addictions (O’Hollaren, 1961), NAD+ infusion therapy is now widely used for the promotion of overall well-being and longevity. Purported benefits of NAD+ IV include, but are not limited to, depression and anxiety reduction, drug and alcohol addiction treatment, hangover relief, fatigue, neurological disorders, athletic performance, and most recently, recovery from symptoms of COVID-19 and post-acute sequelae of SARS-CoV-2 infection (PASC, ‘long-COVID’). Intravenous infusions may be preferred to oral administration under certain clinical circumstances, as direct delivery into the bloodstream can provide 100% bioavailability, which is not achieved through oral supplementation. Nevertheless, despite the wide availability and a broad range of anecdotally reported benefits of NAD+ IV, there is a paucity of human data interrogating its use as a treatment or health-modifying modality.

Aside from the metabolic inefficiency associated with direct exogenous NAD+ administration (via oral, IV, or intramuscular routes) due to the requisite need for its breakdown into its constituent pyridine metabolites before cellular entry, raising extracellular NAD+ (eNAD+) may also provoke maladaptive effects. Under normal physiological conditions, NAD+ is reported to circulate in mammalian extracellular fluids in concentrations between 0.1 and 0.5 μM (Adriouch *et al*., 2012; Gasparrini *et al*., 2021). Increased eNAD+ beyond homeostatic-controlled ranges may represent a pathophysiological trigger, resulting in pro-inflammatory signaling, toxic effects on T-cells in preclinical models, including apoptosis (Adriouch *et al*., 2001; Liu *et al*., 2001), and potentially suppressing immune responses (Liu *et al*., 2001). These findings highlight the need for caution with NAD+ IV, due to its ability to augment eNAD+ to supraphysiologic, potentially pathophysiologic, amounts. Alarmingly, systematic investigations addressing the safety and tolerability of NAD+ IV remain limited despite its widespread use. Clinician- and participant-reported adverse experiences include nausea, diarrhea, muscle cramping, chest pains, and dizziness.

In light of these concerns, the clinical investigation of alternative strategies for boosting NAD through the intravenous route of administration is warranted. Since its recognition as an endogenous form of vitamin B3, as an NAD precursor, nicotinamide riboside (NR) has been the topic of investigation in numerous preclinical and clinical studies (Cantó *et al*., 2012; Samuel A. J. Trammell *et al*., 2016; Airhart *et al*., 2017; Martens *et al*., 2018; Conze *et al*., 2019; Elhassan *et al*., 2019; Brakedal *et al*., 2022; Wu *et al*., 2022). Moreover, the patented form of NR chloride, Niagen®, has received favorable safety reviews from multiple regulatory agencies, including the U.S. Food and Drug Administration (FDA), from which it achieved new dietary ingredient and generally recognized as safe (GRAS) status for use in dietary supplement and food products. Although oral Niagen® supplementation has consistently demonstrated safety and efficacy in dose-dependently augmenting cellular NAD+ levels in human intervention studies at doses up to 3,000 mg/day (Conze *et al*., 2019; Berven *et al*., 2023), its effects following IV administration have yet to be systematically evaluated. Arguably, by bypassing possible gastrointestinal digestive enzyme- and microbiota- mediated degradation and hepatic first-pass metabolism, administering NR intravenously may exert a more potent impact on systemic NAD+ levels in comparison to oral consumption. Additionally, multiple lines of evidence and mechanistic understandings support the premise that intravenous NR (‘NR IV’) offers superior safety and NAD-boosting efficiency over NAD+ IV. Since December 2022, NR chloride has been included on the Bulk Drug Substances Category 1 list under evaluation by FDA as an injectable compound under Section 503B of the Federal Food, Drug, and Cosmetic Act (FDA, 2023).

The aim of the present pilot clinical study is to compare the effects of a single intravenous administration of NAD+, NR, or saline vehicle control, and oral NR supplementation on changes in vitals during and after administration, changes in whole blood NAD+, and tolerable infusion rates. Secondary aims are to compare the clinical chemistry safety profiles of the two approaches of IV supplementation and to identify consumer preferences.

## METHODS

### Ethics & Regulatory Authorization

The study was conducted in accordance with the Declaration of Helsinki and adhered to Good Clinical Practice guidelines. The clinical trial was administered by Nutraceuticals Research Institute at Hopewell Family Care (Hermitage, TN) and received full authorization from Sterling IRB (Protocol Number 23-08-0010), an independent ethics review board which is registered with the National Institutes of Health’s Office for Human Research Protections (OHRP). This authorization remained in effect through the trial and was expanded to include the addition of study 2. The trial was also registered at ClinicalTrials.gov, NCT06382688. All participants provided written informed consent prior to any intervention procedures.

### Trial Design

This was a 2-part study. The first study was a 4-arm randomized, placebo-controlled parallel trial. The 3 IV arms were double-blinded, while the 4th arm involved oral administration making it naturally unblinded. Participants were randomized at a 10:10:10:6 ratio to intravenous placebo (saline), NAD+, NR, or oral NR (oral). The second study was a randomized parallel design trial with two arms for the purpose of evaluating the actual differences in infusion rates in real time. Participants were randomized at a 1:1 ratio to intravenous NR or NAD+.

### Participants

Participants were determined to be eligible for this study if they met the following inclusion criteria: Signed and dated the informed consent form, demonstrated ability to comply with study procedures, live within 100 miles of the Nutraceuticals Research Institute (Franklin, TN) study sites, were 40+ years in age, overall good general health, and females with reproductive potential must have used a highly effective contraception for at least 1 month prior to screening as well as agreeing to use such method during study participation and 1 month after the study end date.

Study one was open to any sex; study two was restricted to biological males.

Participants were not eligible for either study if they met any of the following exclusion criteria: Current diagnosis of any seizure disorder, diabetes or insulin resistance, any kidney or liver disorder, heart disease, cancer, or Parkinson’s Disease, pregnancy, trying to conceive, or breastfeeding, and had any known allergies to any components of the interventions.

### Sample Size (1 P)

The purpose of this study was to identify the continuity and variations between the two approaches to IV administration, to serve as a pilot study to collect data for future research, and to provide data for future powered studies. It was not to establish a statistically significant difference between the two.

A post-hoc power analysis was conducted using G*Power to identify the power of the study. Using the findings from the first study tolerable infusion rate on four groups, with an effect size (F) of 10.12 and a p-value of <.001, the actual power of the study was found to be 99%.

### Randomization & Blinding

Participants were adaptively randomized by age and sex using the method developed by Kang et al. (2008). This ensured even distribution of patients in all groups based on these potentially confounding variables.

Participants and participant-facing study staff were blinded to allocation of interventions, with the exception of the oral group which was naturally unblinded. To maintain blinding, participants were seated in comfortable chairs for IV administration and the IVs were prepared in a different area, out of sight of the participants. The IVs were labeled with the participant’s ID.

### Intervention

The test intervention for both studies was pharmaceutical grade NR chloride (Niagen®), which was obtained from W.R. Grace (South Haven, MI) for preparation from a compounding pharmacy. The first study, which had a total of four groups, included comparison groups of placebo (saline IV), an active comparator (NAD+), and oral administration of NR chloride (Niagen®, ChromaDex). The second study, which had a total of two groups, included the test intervention and active comparator.

Vials of 50 mg/ml of NR or NAD+ were prepared in 10 ml of sterile water by DCA Pharmacy (Franklin, Tennessee) by prescription, for research administration only, and maintained in refrigerated conditions until use. For participants who were randomized to the test group or to the active comparator group, the NR or NAD+ was added to 500 ml of normal saline (B Braun 0.9% sodium chloride injection USP, preservative free) prior to intravenous administration. Those randomly assigned to the saline group received 500ml of normal saline. Participants that were randomly assigned to the oral arm were provided with 500 mg of NR which was taken with water.

In the first study, participants began the infusion at a rate of 20 drops per minute for the first 15 minutes to ensure safety and comfort. After this initial timepoint, participant input was used to increase or decrease the infusion rate in order to meet or maintain the comfort of the participants throughout the duration of the infusion. In the second study, all participants began with the IV line fully open. Participants were closely monitored for the duration of the infusion, and the infusion rate was decreased, as applicable, upon participant request.

Consented participants were instructed to fast for at least 8 hours prior to the time of the infusion and for at least 8 hours prior to the 24-hour assessments (Study 1 only), consuming only water, black coffee, or black tea. In the event of anticipated side effects, such as nausea, upset stomach, or vomiting, participants were offered saltine crackers and ginger during and after the infusions. Participants were provided a standardized lunch (Study 1) after the administration of the test material.

### Outcomes

The safety of the IV infusion was evaluated using a combination of endpoints, including tolerable infusion rate, total number of adverse effects, participant vital signs, a complete blood count (CBC), a comprehensive metabolic panel (CMP), glucose and insulin levels, and the participant’s self-reported subjective experience.

Socioeconomic status control variables included age, race, height, weight, BMI, household income, educational attainment, marital status, and employment status.

Adverse event monitoring began on the first day of the study and continued for 14 days (study one) or 7 days (study two). Participants were instructed to notify the research staff of any new or unusual symptoms during the trial and open-ended questions during data collection solicited unexpected side effects.

### NAD+ Analysis

Finger punctures were conducted using a lancet. Blood was then applied to the circles on the dried blood spot (DBS) card and allowed to dry for a minimum of 3 hours. Dried cards were stored at -20°C until shipment for analysis by LC/MS.

### Statistical Methods

Continuous demographics and randomization success were assessed using two-sample t-tests. Categorical demographic and randomization variables were assessed using chi-square analysis.

Between-groups comparisons of each outcome measured in the CMC and CMP were evaluated using mixed between-within subjects ANOVA with the timepoint being the within subjects’ factor and the group assignment as the between subjects factor. Mauchly’s test of sphericity was used to confirm assumptions. Violations of the assumption of sphericity are addressed with the Greenhouse-Geisser correction.

To assess vital signs and subdomains on the sleep and energy scales, between-groups comparisons were assessed using analysis of covariance (ANCOVA) with baseline scores as the covariate and a Bonferroni correction. Levene’s test was used to check and confirm the assumption of homogeneity of variances.

Participants were analyzed using an intent-to-treat (ITT) analysis. Those who were randomized to a group and began the study were evaluated in the statistical analysis. All data were analyzed using STATA v17.

### Additional Analyses

Additional analyses were conducted using GraphPad Prism (Ver. 10.0.2). Statistical significance was determined as p<0.05, with comparisons of the test articles compared to saline or between group comparisons. 2way ANOVA with Tukey’s multiple comparisons test with a single pooled variance was used to determine significance in hematology, clinical chemistry, and vitals. Time in chair was analyzed for study 1 by a one-way ANOVA with multiple comparisons, study 2 was analyzed by unpaired t-test. The baseline variable of age was analyzed for differences by one way ANOVA with multiple comparisons for study.

## RESULTS

### Participant Flow

A total of 45 individuals were assessed for the first study. Of these participants, 2 did not provide consent, and a total of 43 qualified and provided informed consent. Of these, 6 withdrew due to scheduling conflicts. A total of 37 were randomized into one of the four groups and received the intervention, NAD+ IV (n=10); NR IV (n=11); saline (n=6); oral NR (n=10). For the second study, a total of 22 individuals were assessed; 16 qualified and provided informed consent, were randomized to a group, and received an intervention; NAD+ IV (n=8); NR IV (n=8). No participants were removed from the study or withdrew from either study once the trial began. All participants in both studies provided written informed consent prior to any intervention related procedures. Participants were followed until 14 days (first study) or 7 days (second study) after the trial for reporting of adverse events. See Flow Chart (**Figure 1**).

### Baseline Data

Baseline descriptive statistics were evaluated, and t-tests were performed to ensure balance between each group. No differences between the two groups were identified, indicating that randomization successfully balanced the groups on known factors. In the first study, participants were mostly white (89%) and 41 years old (27%). Gender was indicated by their sex assigned at birth. The gender ratio for this study consisted of 59% male and 41% female. In the second study, all participants were male, with an average age of 45.57 years (range: 40-64). Health history was also similar between groups. There were no differences between groups on any of the baseline control variables. However, these results should be interpreted with caution due to the small sample sizes (**Table 1**).

**Table 1.** Study 1 and 2 Participant Demographics & Baseline Vitals.

| Study | Characteristic | Results (Mean ± SD) | | | |
| --- | --- | --- | --- | --- | --- |
|  |  | NAD+ IV | NR IV | Saline | Oral |
| Study 1 | Sex (M/F) | 7/3 | 7/4 | 3/3 | 5/5 |
|  | Age Mean years (min, max) | 41.60 (40, 46) | 41.50 (40, 45) | 44.00 (40, 51) | 44.20 (40, 48) |
|  | Race | White: 7<br>Hispanic/Latino: 2<br>Black/African-American: 1 | White: 11 | White: 6 | White: 9<br>Filipino: 1 |
| | BMI | 25.55 ± 5.38 | 25.35 ± 3.01 | 28.60 ± 4.71 | 26.90 ± 3.02 |
| | Baseline Systolic BP | 118.2 ± 12.57 | 120.7 ± 16.51 | 121.5 ± 14.50 | 113.8 ± 12.40 |
| | Baseline Diastolic BP | 78.60 ± 12.66 | 75.27 ± 10.86<sup>∞</sup> | 74.17 ± 9.020 | 73.11 ± 8.796<sup>∞</sup> |
| | Baseline Pulse | 73.00 ± 7.513 | 68.09 ± 11.58 | 77.67 ± 12.69 | 66.44 ± 9.126* |
| Study 2 | Sex (M/F) | 8/0 | 8/0 | N/A | N/A |
|  | Age Mean years (min, max) | 47.29 (41, 64) | 43.88 (40, 52) | N/A | N/A |
|  | Race | White: 7<br>Other: 1 | White: 6<br>Other: 2 | N/A | N/A |

∞ indicates between group significance. \* indicates significance compared to saline. ANOVA p<0.05

### Intent to Treat

For the first study, data from 37 patients were available for intent to treat analysis; for the second study, data from 16 patients were available for intent to treat analysis. **Table 2** describes the timepoints for the various assessments that were conducted.

**Table 2.** Study Design.

| Study | Procedures | Day 0 | Day 1 Baseline | Day 1 Post Infusion | Day 1 3 hr Post Infusion | Day 1 6 hr Post Infusion | Day 2 24 hr Post Infusion | Day 7 | Day 14 |
| --- | --- | --- | --- | --- | --- | --- | --- | --- | --- |
| | | | | (± 10 minutes) | (± 15 min) | (± 45 min) | (± 3 hours) | (± 1 day) | (± 1 day) |
| Study 1 | Screening | X |  |  |  |  |  |  |  |
|  | Informed Consent | X | X |  |  |  |  |  |  |
|  | Demographics & Randomization | X |  |  |  |  |  |  |  |
|  | Administer IV Infusion/Oral NR |  | X |  |  |  |  |  |  |
|  | Vitals |  | X | X | X | X | X |  |  |
|  | Blood Draw (CMP, CBC, hematology) |  | X |  | X |  | X |  |  |
|  | Dried Blood Spot Collection |  | X | X | X | X | X | X | X |
|  | Subjective Experience |  |  | X | X | X | X | X | X |
|  | Adverse event reporting |  | X | X | X | X | X | X | X |
| Study 2 | Screening | X |  |  |  |  |  |  |  |
|  | Informed Consent | X | X |  |  |  |  |  |  |
|  | Demographics & Randomization | X |  |  |  |  |  |  |  |
|  | Administer IV Infusion/Oral NR |  | X |  |  |  |  |  |  |
|  | Vitals |  | X | X |  |  |  |  |  |
|  | Subjective Experience |  |  | X |  |  |  |  |  |
|  | Adverse event review and evaluation |  | X | X |  |  |  | X |  |

**Table 3.**
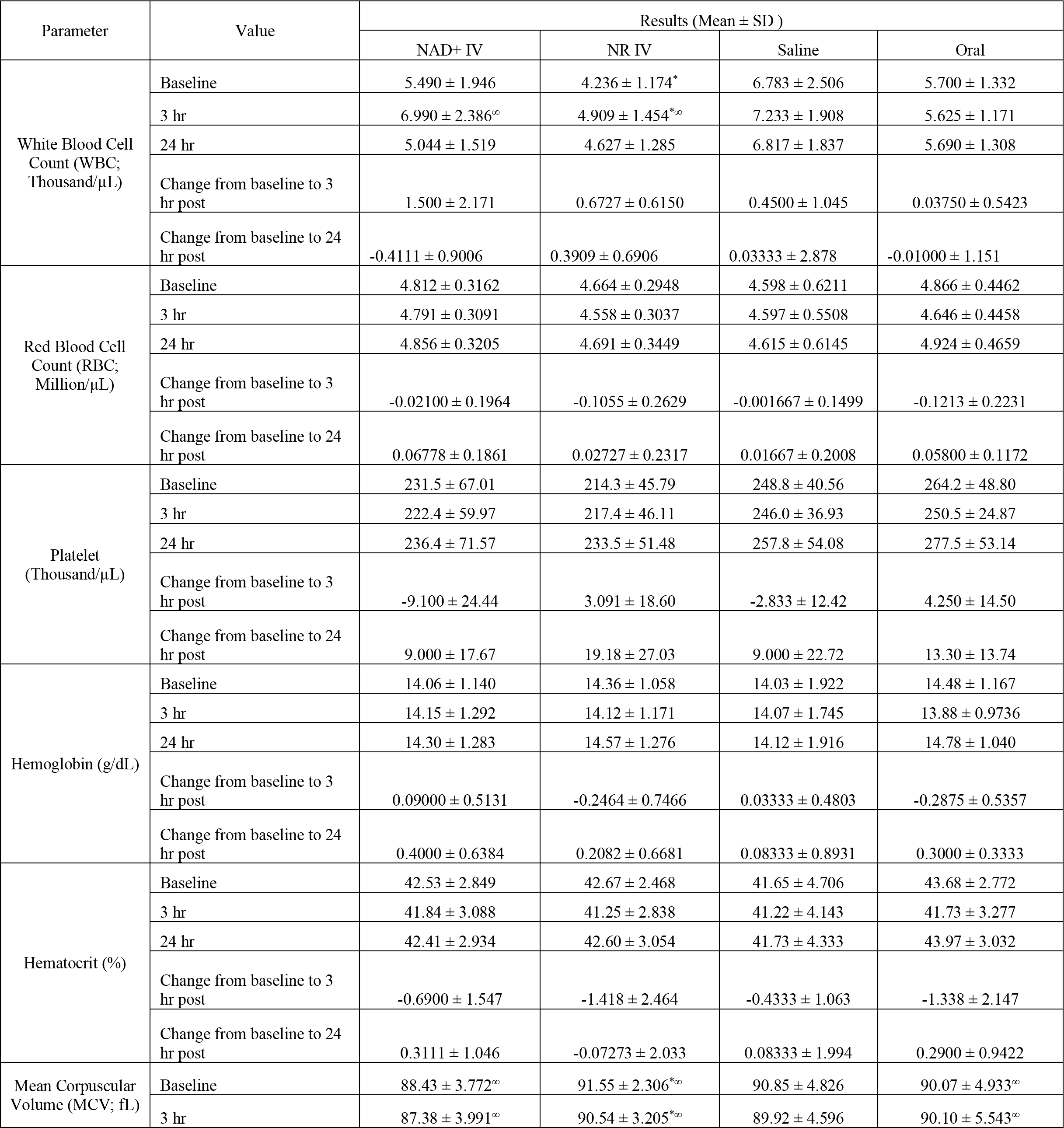
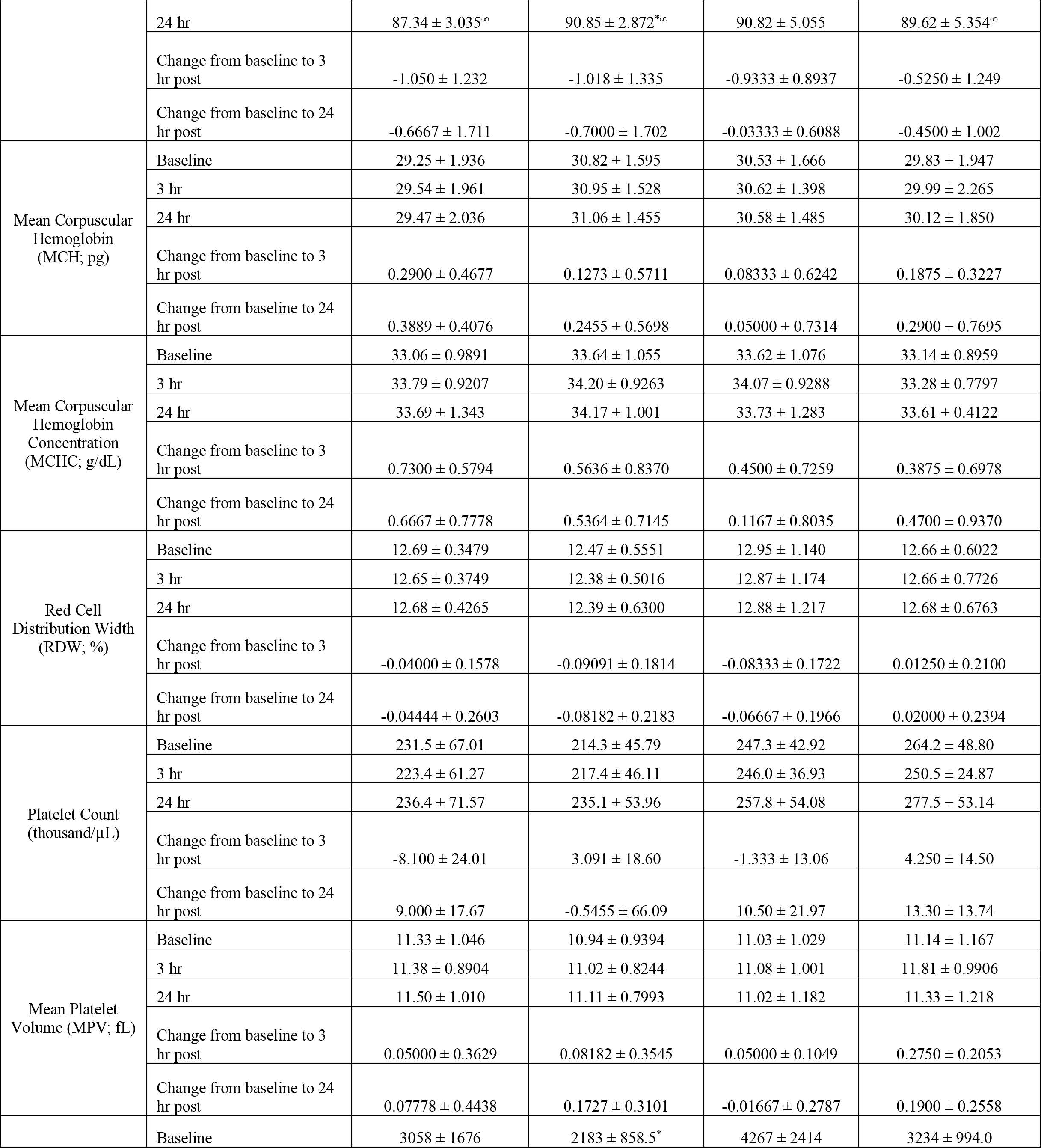
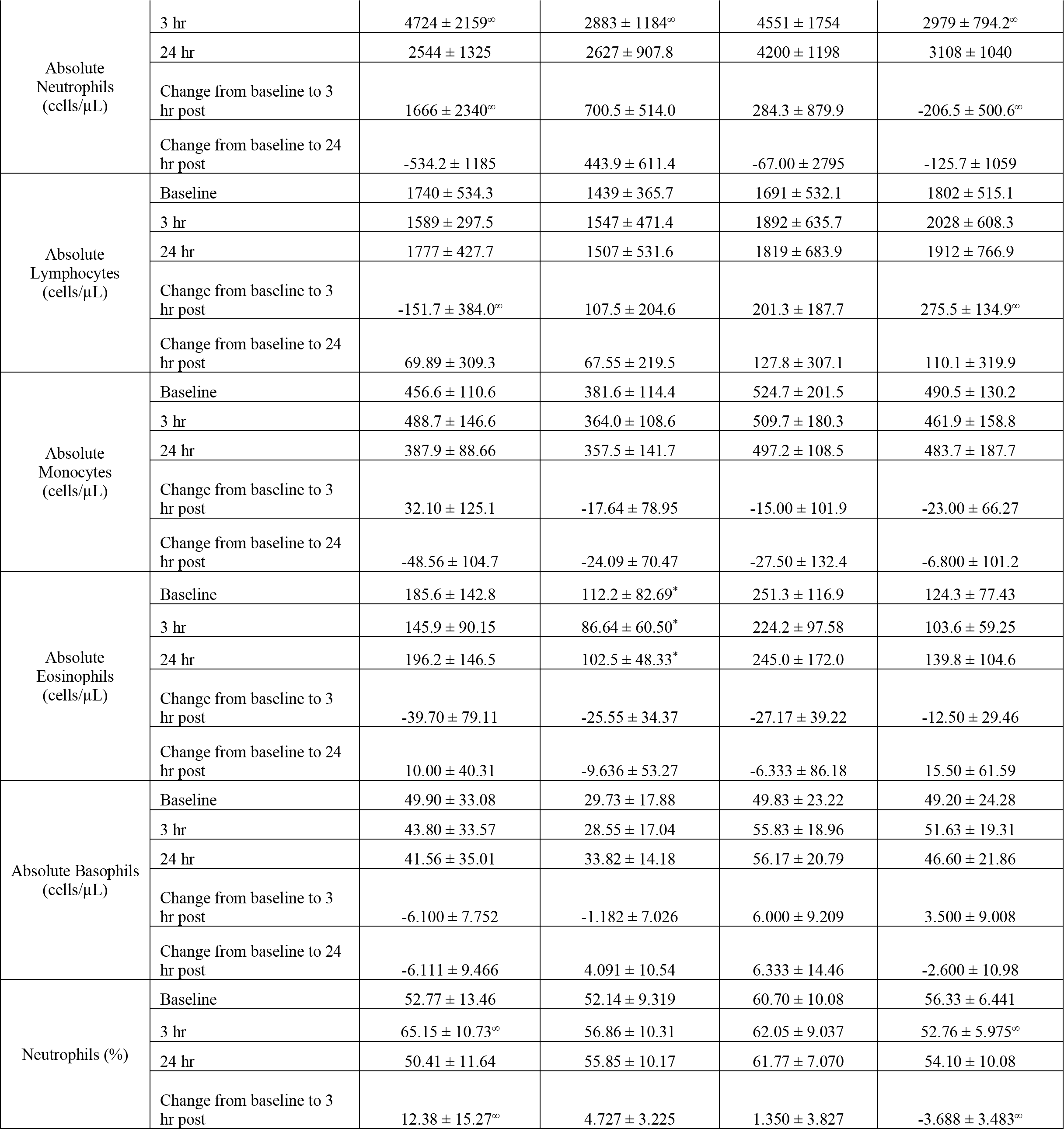
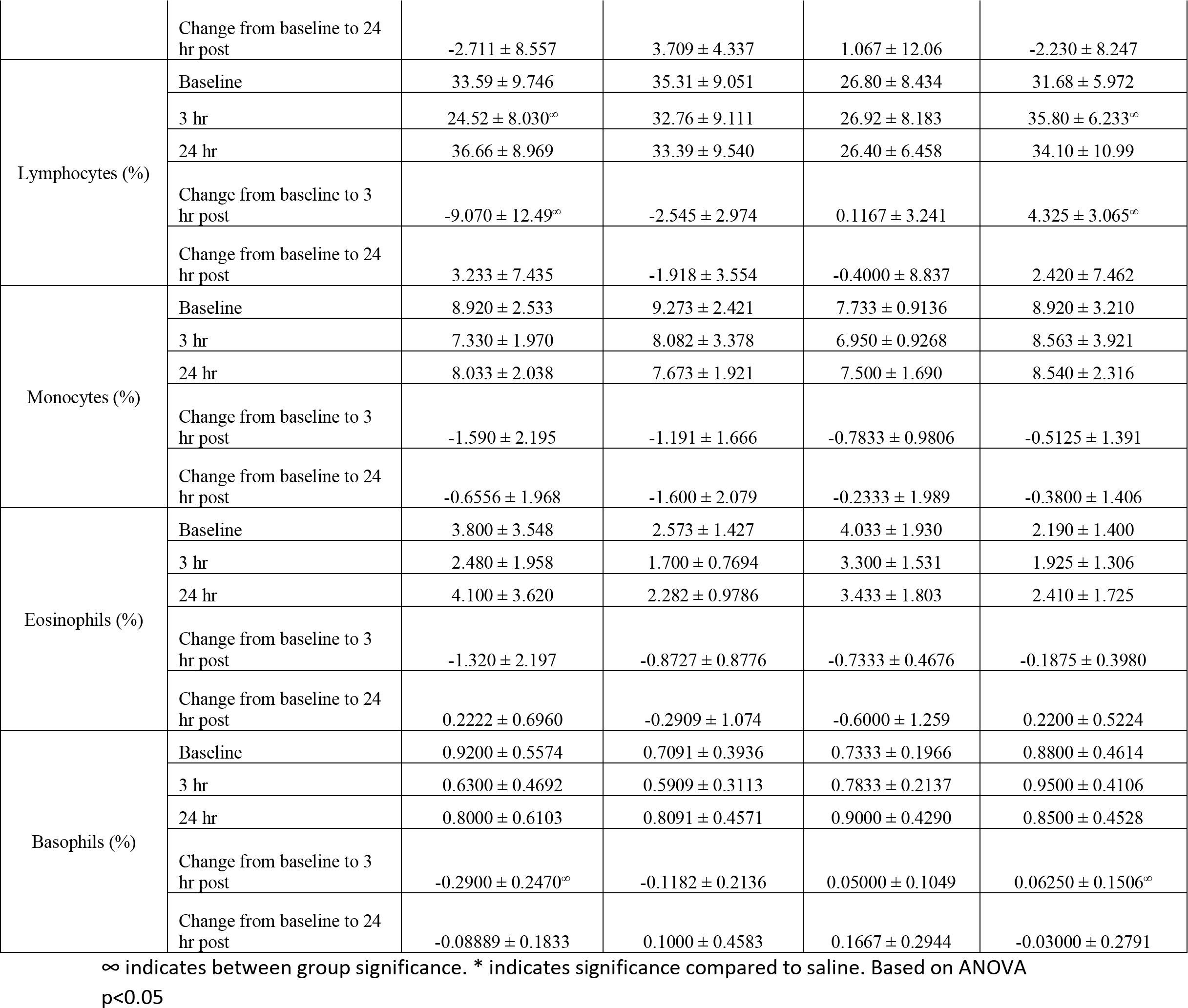
Study 1 Complete Blood Count (hematology)

**Table 4.**
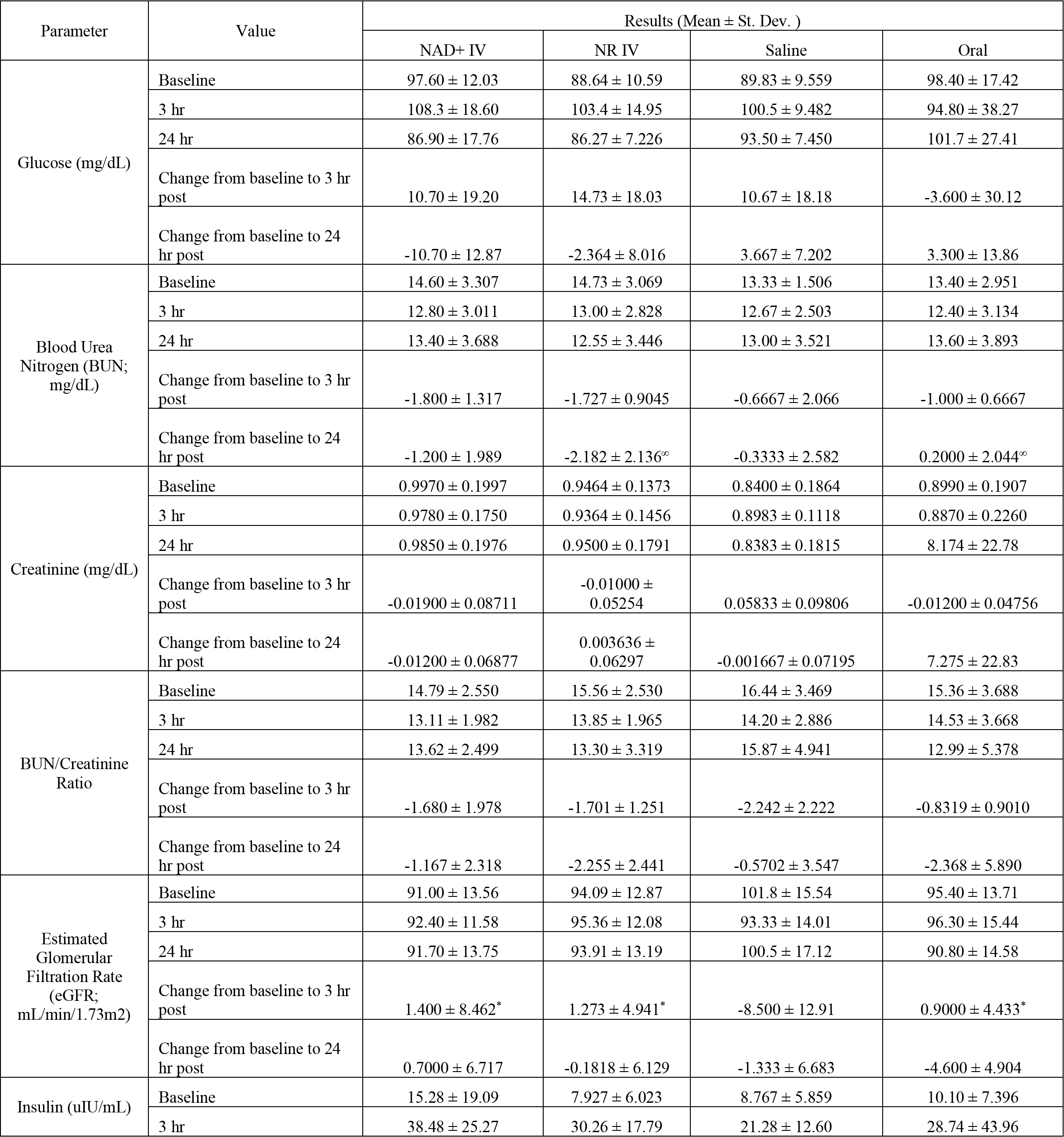
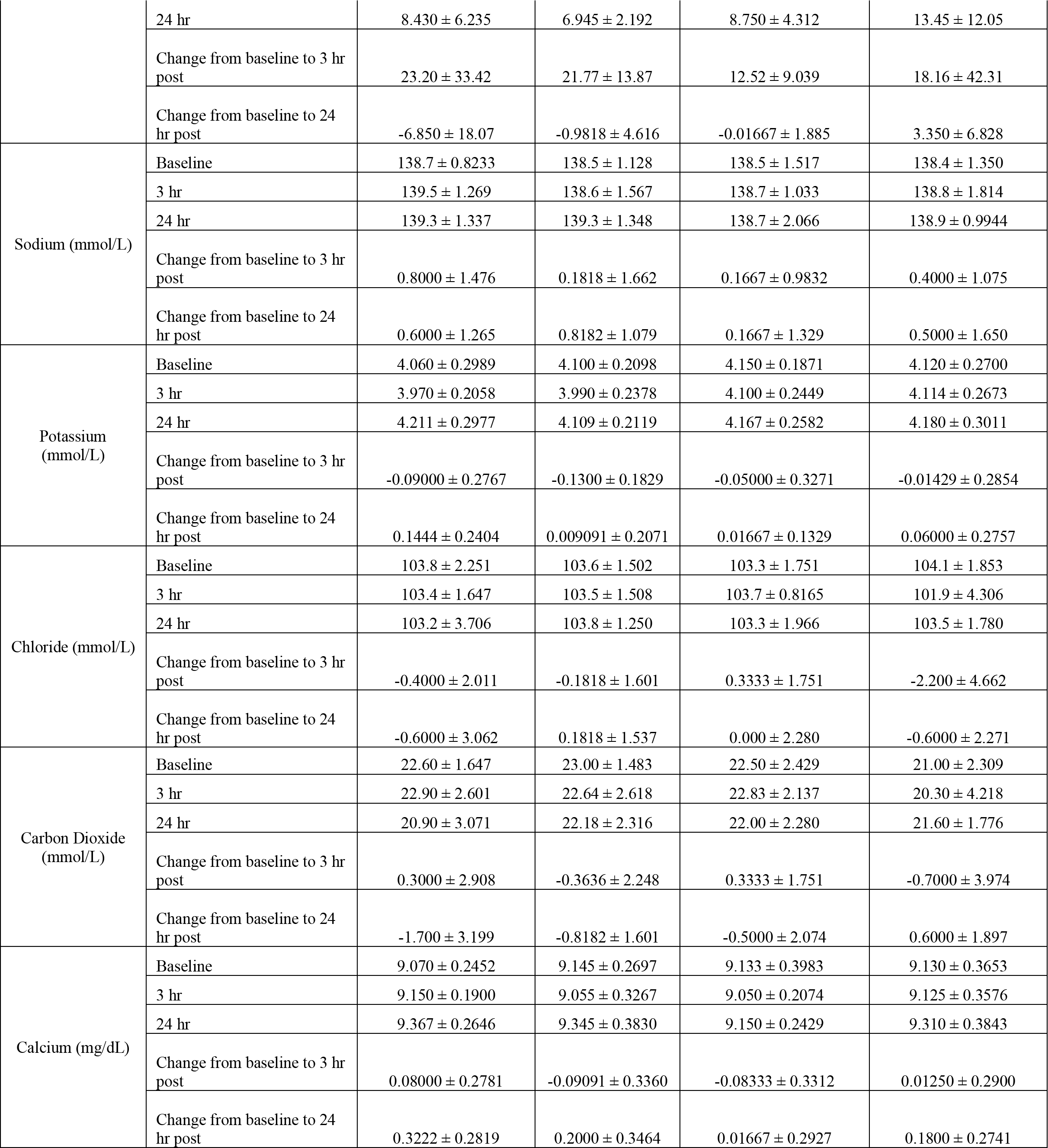
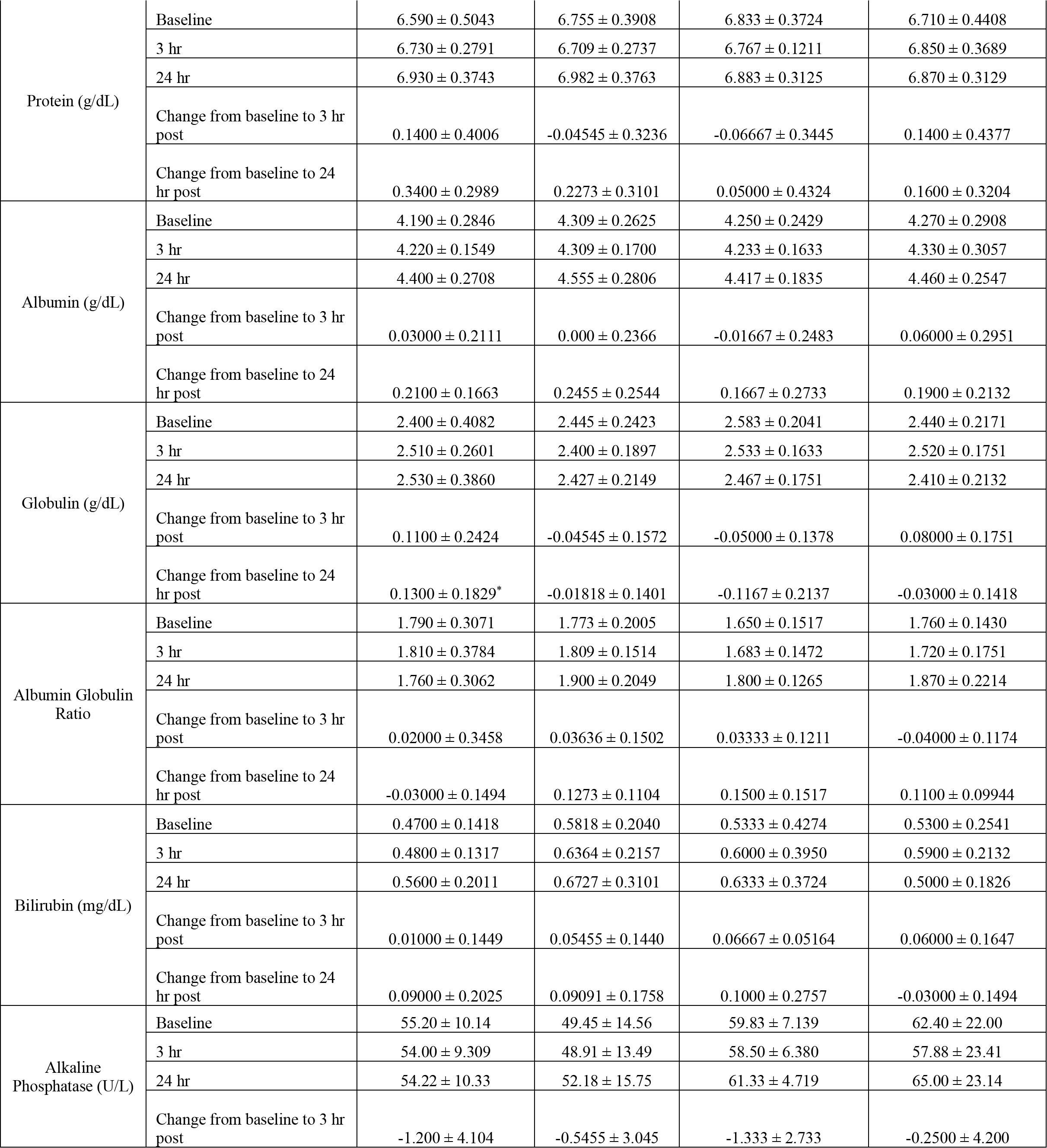
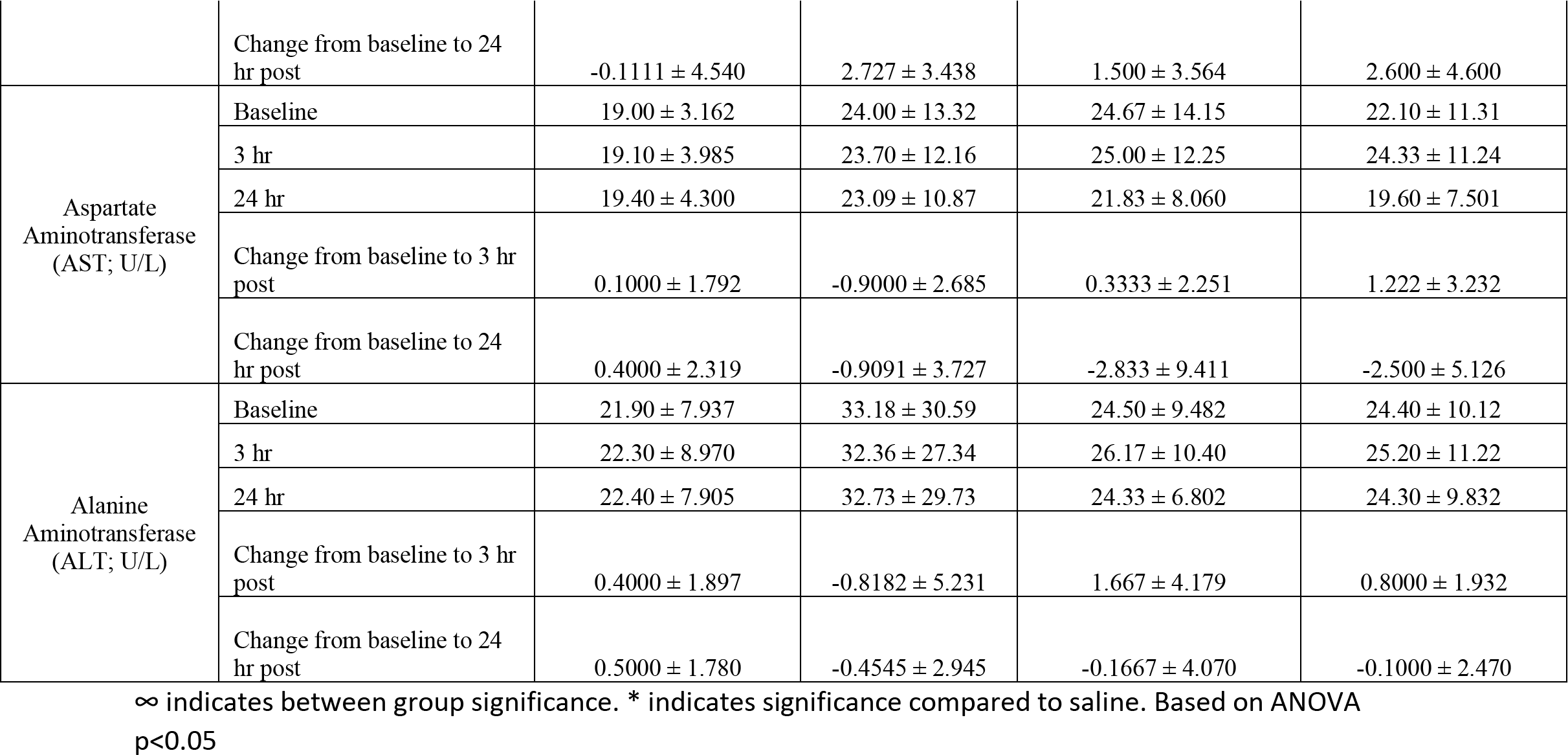
Study 1 Clinical Chemistry.

### Vital Signs

Post-intervention systolic blood pressure (mmHg) values were 117.20 (SD:14.54), 121.40 (SD:13.84, 115.50 (SD:4.23), and 108.56 (SD:9.08) in the NAD+ IV, NR IV, saline, and oral groups, respectively. An ANCOVA with baseline systolic blood pressure as the covariate revealed there were no significant changes between groups on scores (F (1,34) = 0.22, p=0.882).

Post-intervention diastolic blood pressure (mmHg) values were 73.40 (SD:13.15), 79.60 (SD:6.24), 74.83 (SD:6.59), and 71.00 (SD:6.24) in the NAD+ IV, NR IV, saline, and oral groups, respectively. An ANCOVA with baseline systolic blood pressure as the covariate revealed there were no significant changes between groups on scores (F (1,34) = 1.61, p=0.21).

Post-intervention heart rate (beats per minute, BPM) values were 68.40 (SD:8.10), 63.45 (SD:9.42), 65.83 (SD:12.69), and 65.78 (SD:7.95) in the NAD+ IV, NR IV, saline, and oral groups, respectively. An ANCOVA with baseline systolic blood pressure as the covariate revealed there were no significant changes between groups on scores (F (1,34) = 2.15, p=0.143).

### Tolerable Infusion Rates

In the first study, NR IV infusion intake varied from 1 hour 10 minutes to 3 hours 7 minutes, with the average being 2 hours and 7 minutes (127.2±40.82 minutes). In contrast, the NAD+ IV infusion intake varied from 1 hour and 47 minutes to 4 hours 32 minutes, with the average being 3 hours and 3 minutes (182.9±55.93 minutes). The saline group ranged from 1 hour and 10 minutes to 1 hour and 39 minutes, with the average being 1 hour and 25 minutes (85.17±9.745 minutes). A one-way analysis of variance identified a significant difference between groups (F (1,26) = 10.12, p=<.001). A Bonferroni post-hoc test identified significant differences between the NAD+ IV and NR groups (t=-2.94, p=.022) and between the NAD+ IV and saline groups (t=- 4.36, p=.001). The effect size, calculated as partial eta squared, was 0.457, which far exceeds the classification of large described by Cohen et al. To further elucidate the variation in total infusion time between NR IV and NAD+ IV, in the second study an additional 16 male patients received an IV injection of each substance at a 1:1 ratio (n=16). NR IV infusions ranged from 11 minutes to 41 minutes, while NAD+ IV infusions ranged from 20 minutes to 184 minutes. The average infusion rate for NR IV was 24.75 (SD: 8.33) minutes, compared to an average infusion rate for NAD+ IV of 98.88 (SD: 46.70) minutes. A two-tailed t-test was used to compare the differences and revealed that the NR IV infusion rate was significantly and substantially lower than the NAD+ IV infusion rate (t(14)=-4.42, p=<.001), resulting in a 75% decrease in total time required for NR IV compared to NAD+ IV.

### Patient Subjective Experience

Among the NR IV patients, the most commonly reported sensation was tingling in the mouth and in the extremities. This was described by patients as “tingly,” “slightly painful,” “burning,” and “weird.” One described the feeling “like eating pop rocks.” Patients also reported feeling a sensation of pressure in the head and ears to the nursing staff. They described it as feeling congested and during the IV had a “runny nose” at times.

By contrast, the majority of NAD+ IV patients described their comfort level as “low.” They self-reported anxiety, headaches, nausea, and sudden urges to have a bowel movement or diarrhea.

They described it as feeling “chest tightness and a little woozy,” “hot flashes,” “feeling queasy,” and “cramping in my stomach.”

Nursing staff reported additional symptom descriptors including “feeling gassy,” “muscle weakness,” “unsettled stomach,” and “stomach cramping.” Approximately half of the patients receiving NAD+ IV had a bowel movement during the IV.

### Chemical Chemistry

Blood chemistry remained relatively stable during the intervention period. A mixed between-within subjects analysis of variance was conducted to assess potential variations between groups and timepoints. While some significant timepoint effects were observed, no significant between groups effects were observed on the endpoints obtained in the CMC and CMP tests. Significant between-groups differences were identified for the outcomes of glucose and insulin.

For the outcome of glucose, the overall model was significant (Wald χ²(11, n=37)=41.76, p=<.001). Post hoc tests identified significant differences between the NAD+ IV group and the saline group (χ²=7.61; p=.022) and the oral group (χ²=18.42; p=<.001). For the outcome of insulin, the overall model was significant (Wald χ²(11, n=37)=56.30, p=<.001), but there were no significant between-groups differences identified.

### Additional Analyses

While there were statistically significant differences between saline, the test group (NR IV) and active comparator group (NAD+ IV), the changes were not deemed clinically significant. An ANOVA analysis indicated a significant difference for NAD+ IV and NR IV to saline in the estimated glomerular filtration rate change from baseline to 3 hours post infusion (NAD+ IV: 1.400 ± 8.462; p=0.0341, NR IV: 1.273 ± 4.941; p=0.0330, Saline: -8.500 ± 12.91). This change was not deemed clinically significant. The change in globulin from baseline to 24 hours post infusion was significantly different in NAD+ IV compared to saline (0.1300 ± 0.1829 and - 0.1167 ± 0.2137 respectively; p=0.0418).

An ANOVA analysis indicated a between group significance at 3 hours post infusion in absolute neutrophils between NAD+ IV and NR IV (4724 ± 2159 and 2883 ± 1184 respectively; p=0.0164), this also shows a clinical significance with the changes in NAD+ IV likely due to inflammation. Baseline absolute neutrophil levels were statistically different in the NR IV group compared to the saline group (2183 ± 858.5; p=0.0203). Absolute eosinophils were significantly different in the NR IV group at all three time points compared to saline (baseline: 112.2 ± 82.69; p=0.0413, 3 hours post infusion: 86.64 ± 60.50; p=0.0448, 24 hours post infusion: 102.5 ± 48.33; p=0.0350). The mean corpuscular volume was significantly different at all three time points for NR IV compared to the saline group (baseline: 91.55 ± 2.306 and 90.85 ± 4.826 respectively; p<0.0001, 3 hours post infusion: 90.54 ± 3.205 and 89.92 ± 4.596 respectively; p<0.0001, 24 hours post infusion: 90.85 ± 2.872 and 90.82 ± 5.055 respectively; p<0.0001). MCV was also significantly different between NAD+ IV and NR IV at all three time points (baseline: 88.43 ± 3.772 and 91.55 ± 2.306 respectively; p<0.0001, 3 hours post infusion: 87.38 ± 3.991 and 90.54 ± 3.205 respectively; p<0.0001, 24 hours post infusion: 87.34 ± 3.035 and 90.85 ± 2.872 respectively; p<0.0001). White blood cell count showed statistically significant differences in the NR IV group compared to saline at baseline (4.236 ± 1.174 and 6.783 ± 2.506 respectively; p=0.0161) and 3 hours post infusion (4.909 ± 1.454 and 7.233 ± 1.908 respectively; p=0.0336). Between group differences were observed at 3 hours post infusion between NAD+ IV and NR IV (6.990 ± 2.386 and 4.909 ± 1.454 respectively; p=0.0249) though these were not deemed clinically significant.

### Adverse Events

Participants were monitored for adverse events and for the development of any exclusion criteria during the intervention. One participant in the NR IV group stumbled on a low corner of a bookshelf while engaging in physical activity in the waiting area. This resulted in a small cut, which was treated with a Band-Aid. This AE was classified as *mild* and was determined to be not related to the intervention. No other AEs were identified during the intervention period or the follow-up period.

### NAD+ Analysis

The use of dried blood spots (DBS) allowed for the analysis of NAD+ levels based upon samples that could be prepared at the clinic or by the study participants from home. While 36 of 37 participants in study 1 utilized the DBS at all time points, not all samples were analyzable. For the baseline DBS collected for NAD+ IV, NR IV, saline, and oral NR, the usable samples were (n= 6,6,4,8), at t=10 minutes after the completion of the infusion (n=9,7,6,7), at t=3 hours (n=8,9,6,8), at t-6 hours (n=9,9,6,10); at t=24 hours (n=8,8,5,8); t=7 days (n=7,8,5,8) and t=14 days (n=7,9,5,9), respectively (**Figure 4**). NAD+ IV does not appear to increase whole blood NAD+ until 24 hours, with a 2%, average increase, followed by increases at 7 and 14 days, to 8.8 and 15.1% increases, respectively, relative to baseline. For NR IV, NAD+ levels appeared to peak at 3 hours with a 20.7% increase compared to baseline, which was decreased to 16%, 8.1%, and 5.5% at 24 hours, 7- and 14-days, respectively. When comparing the between-group analyses, there were not statistically significant differences (ANOVA) at baseline, 24-hours (p=.242; R2=.151), 7-days (p=.949; R2=.014), or 14-days (p=.420; R2=.101). However, the model was significant for the 3-hour timepoint (p=.010; R2=.337), as significant differences were observed between the NAD+ and NR IV group (t=2.81, p=.009), between the NR-IV and saline groups (t= -2.63, p=.014), and between the NR IV and oral NR groups (t= -3.27, p=.003) and at 6-hours (p=.032; R2=.251). Thus, NR IV resulted in a statistically significant increase in whole blood NAD+ at 3 hours, relative to the placebo and NAD+ IV groups. At 6 hours, near-significant differences were observed between the NAD+ and NR IV group (t= 1.99, p=.055), and significant differences were observed between the NR IV and saline groups (t= -2.80, p=.009), and between the NR IV and oral NR groups (t= -2.59, p=.015).

**Figure 3.**
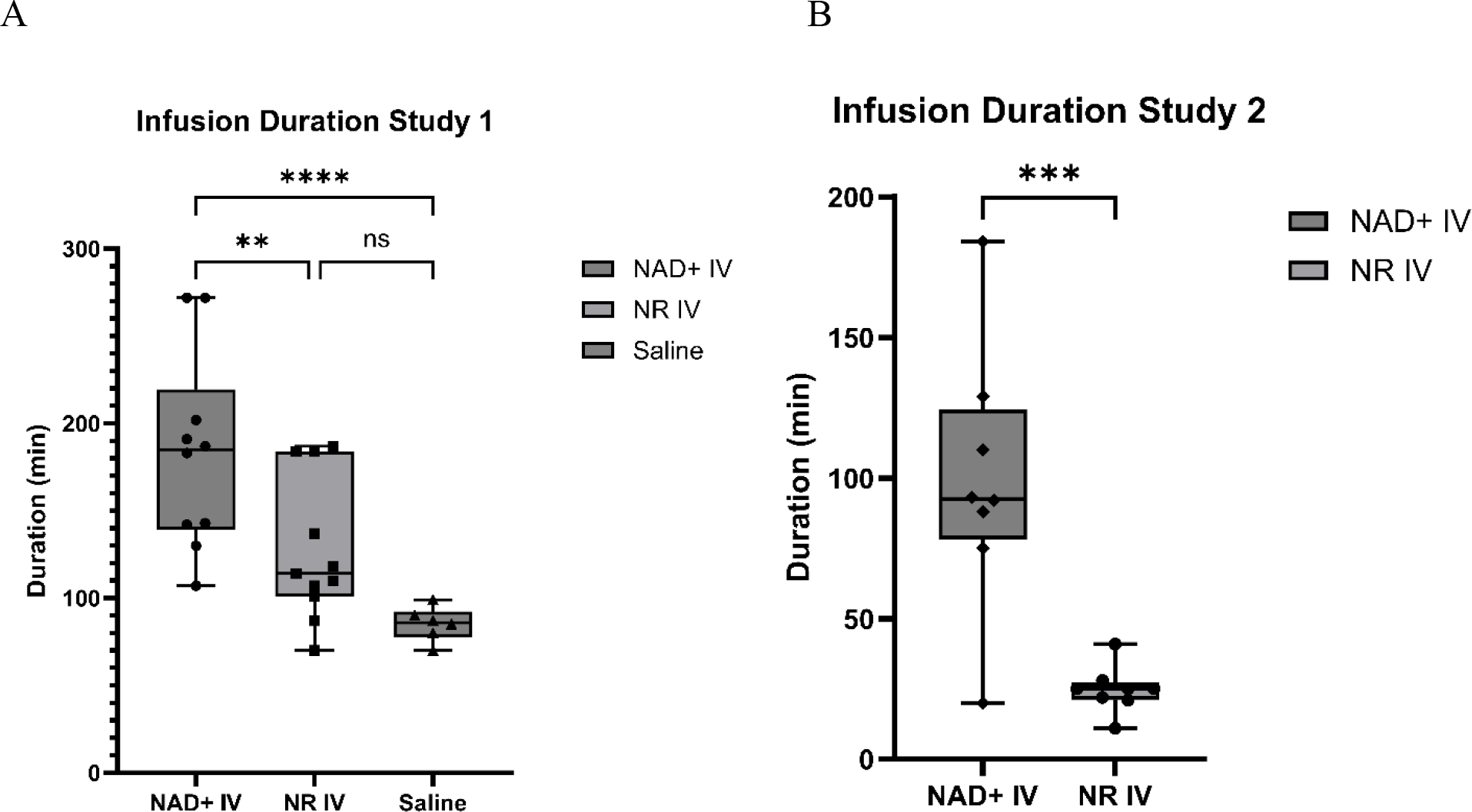
Study 1 & 2 Infusion Duration. A) Infusion duration of Study 1 in minutes, line at the median and whiskers to the minimum and maximum values. B) Infusion duration of Study 2 in minutes, line at the median and whiskers to the minimum and maximum values. Analysis by ANOVA; ns – not significant *p<0.05, **p<0.01, *** p<0.001, **** p<0.0001

**Figure 4.**
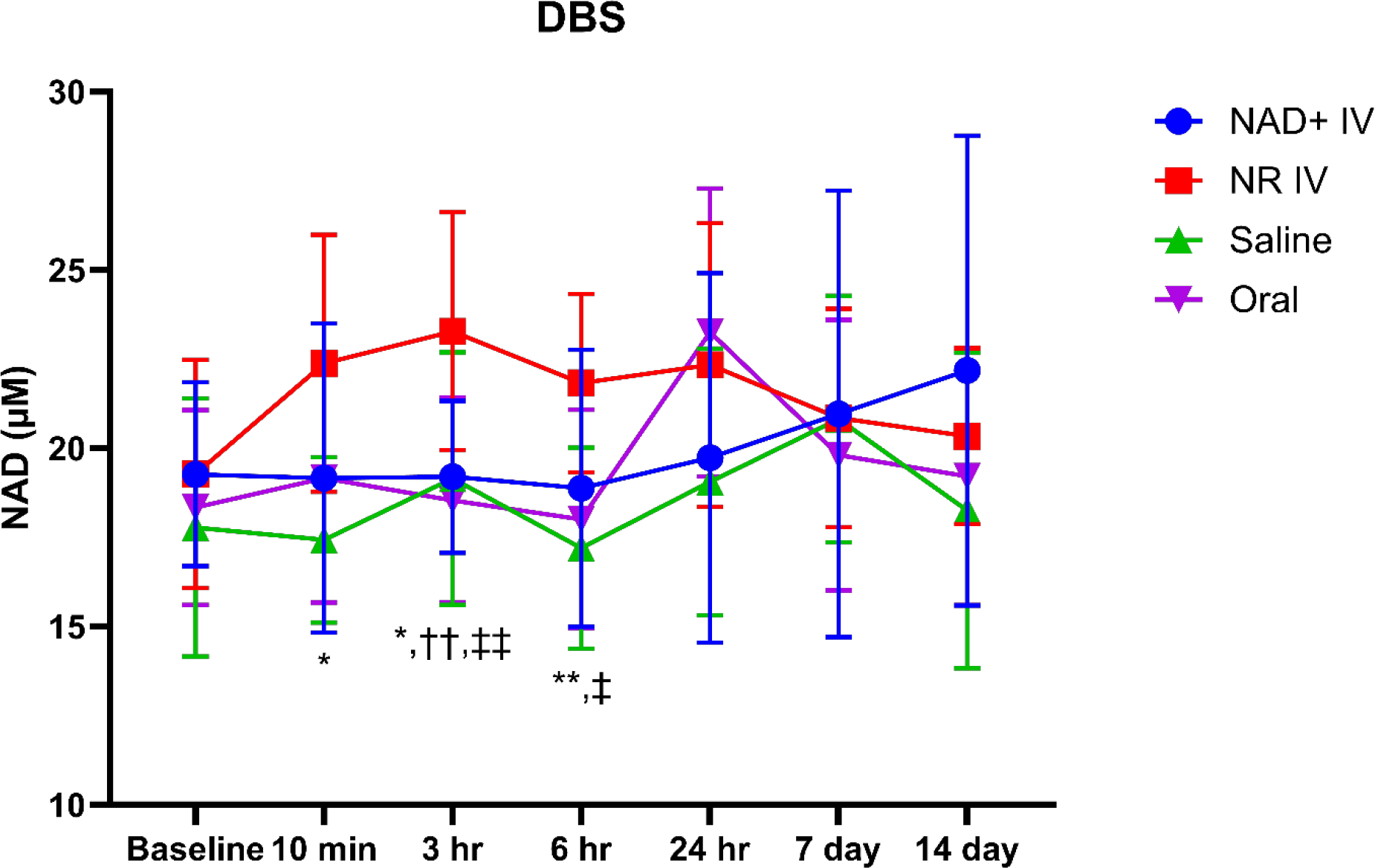
NAD measurements as assessed by dried blood spots. Dried blood spots were collected at various time points to observe the kinetics of NAD+ changes following interventions with NAD+ IV, NR IV, saline, or oral NR. NAD levels are reported as means with standard deviation. Analysis by ANOVA; *, ** = NR IV vs Saline (*p<0.1, **p<0.01); †, †† = NAD vs NR IV (††p<0.01); ‡, ‡‡ = NR IV vs oral (‡p<0.1, ‡‡p<0.01)

## DISCUSSION

NAD+ is an essential coenzyme required for cellular functions and health maintenance. During aging, the accumulation of metabolic stressors that activate NAD+ consuming enzymes, including CD38 and poly (ADP-ribose) polymerases (PARPs) likely result in or contribute to the decline in NAD+ availability (McReynolds *et al*., 2020). While the definition of ‘normal’ NAD+ levels in blood and tissue has yet to be determined scientifically, it is broadly recognized that the maintenance of sufficient intracellular NAD+ pools is required for optimal health.

The clinical use of intravenous NAD+ in the United States was popularized by Paula Norris Mestayer and Dr. Richard Mastayer of the Springfield Wellness Center in the early 2000s and is now offered at clinics throughout the world to treat specific addictions and neurological conditions, as well as general wellness support. Despite its popularity, the science behind the safety and efficacy of NAD+ IV is minimal. Moreover, on a mechanistic basis, there is concern that the presence of increased eNAD+ following NAD+ IV administration may be recognized as a pathophysiological signal by the immune system, resulting in an inflammatory response (Adriouch *et al*., 2012; Audrito *et al*., 2021). Anecdotally, the infusions have been described as painful or uncomfortable, causing gastrointestinal disturbances, thereby necessitating an extremely slow rate of administration. Consistent with these reports, Grant et al. 2019 required an intravenous infusion rate of 2 mg/min over six hours to administer 750 mg of NAD+ to participants without adverse events. The requirement of a six-hour infusion time underscores the time inefficiency of such a method (Grant *et al*., 2019). In the present study, the tolerance to intravenous NAD+ and NR was variable and individual. The majority of participants reported adverse experiences during the NAD+ IV infusion, e.g., nausea, headache, diarrhea, and muscle tightness, in contrast to reports of tingling and slight nausea and coldness in the NR IV group. In both groups, symptoms were resolved once the infusion was completed. In study 1, the infusion rates were normalized for all participants at the beginning of the infusion to ensure safety, as infusion rates and related side effects have not previously been documented in the literature.

Due to the nature of the infusion administration in study 1, study 2 was employed to create a true head-to-head comparison of the infusion tolerance and rate between NAD+ IV and NR IV, as general safety was demonstrated in study 1. As such, it was determined that the average infusion time for NR IV was ¼ of the time for NAD+ IV.

To evaluate the clinical safety of the infusions, the first study examined changes in laboratory metrics relative to the saline control group, 500 mg of NR in the test group, 500 mg of NAD+ as an active comparator, and 500 mg of oral Niagen for bridging the effects of oral and IV administration of NR. The NR IV, saline, and oral group participants did not display any clinically relevant changes in the comprehensive metabolic panel, which included BUN, creatinine, sodium, potassium, calcium, CO2, AST, ALT, Alk Phos, protein, and albumin, or in the CBC with differential. These results are generally consistent with the observations made by Grant et al. in a study investigating the effects of a 6-hour NAD+ IV infusion (Grant et al. 2019). Similarly, intravenous administration of NR was also well-tolerated with no clinically relevant changes in laboratory markers. Unlike Grant et al. 2019, however, who reported statistically significant (though not clinically relevant) changes in circulating bilirubin and AST levels 8 hours after NAD+ IV initiation, we failed to detect significant differences in either indicator of liver function at the 3- and 24-hour timepoints in the NAD+ IV group. These apparent discordances may potentially be explained by the fact that the blood samples for clinical chemistry measurements were collected at different timepoints, as well as differences in dose utilized between the two studies. Likewise, consistent with findings from previously published clinical studies, oral Niagen ingestion was safe and well tolerated.

Noteworthy were shifts in the upward trends for both glucose and insulin concentrations in all four participant groups, including the controls. These parameters were likely influenced by the consumption of food before the 3-hour assessment, which represented a deviation from the original protocol. Thus, this precludes us from ascertaining the actual effects of the interventions on these blood parameters.

Concerning changes in hematological parameters, the NAD+ IV group notably presented with clinically significant elevations in white blood cells and increases in absolute and percentage neutrophils. The increase in neutrophils from baseline to three-hours post-infusion are commonly attributed to inflammatory responses, physiological or psychological stressors, including immunological responses (Tahir and Zahra, 2023). Therefore, these observations are consistent with the notion that NAD+ IV elevates eNAD+ such that it is interpreted by the immune system as a pathological event (Audrito *et al*., 2021). These changes were not observed in the saline, NR IV group, nor in the oral group.

While hematology and clinical chemistry assessments were not incorporated as part of the inclusion criteria, it is worth noting that the mean CO2 levels were lower than typically seen in clinical practice across all groups; two participants had elevated AST/ALT at baseline and two participants were borderline anemic. Additionally, one participant’s glucose and insulin levels at baseline and 24 hours suggest that the fasting protocol was not adhered to, and it is likely that a carbohydrate-rich meal was consumed prior to the test. Additionally, the neutrophil responses for this individual at the three-hour mark suggest that the participant may have been either immunologically dealing with an infection, stress response or other inflammatory response.

The results of the present study are broadly in line with the work of Kimura and colleagues, who, in a single-arm open-label study reported that the intravenous administration of another NAD+ precursor, nicotinamide mononucleotide (NMN, 300 mg dissolved in 100 mL saline, infused at a rate of 5 mL/min) was safe and well-tolerated in 10 healthy Japanese adults without evidence of untoward effects on organ function (Kimura *et al*., 2022). Further, the investigators found that intravenous NMN elevated blood NAD+ levels at multiple timepoints relative to baseline.

Similar to NAD+, NMN is a phosphorylated compound that requires extracellular dephosphorylation to form NR or nicotinamide, which are readily taken up by the cell to generate NAD+ (Ratajczak *et al*., 2016; Anthony A Sauve *et al*., 2023). Therefore, NR is proposed to represent the more efficient means of boosting the intracellular NAD+ pool, and the health benefits of NMN administration are likely mediated through its requisite extracellular dephosphorylation to NR.

Oral supplementation with NAD+ precursors has increased in popularity over the past decade, as a strategy to support healthy aging. NAD+ is directly or indirectly involved with each of the molecular hallmarks of aging, which describe the cellular mechanisms of the aging processes (López-Otín *et al*., 2023). The use of oral supplementation with NR has become a strategy for elevating NAD+ to support healthy aging. Similarly, the use of intravenous methods to boost NAD+ is increasing in popularity, though the evidence of safety and effectiveness of NAD+ IV has been minimal in the peer-reviewed literature. To date, Ross et al. 2019 has shown an increase in NAD+ in the plasma after NAD+ IV infusion, but prior to this article, such information pertaining to the changes in whole blood have not been described. In this study, NAD+ IV did not significantly elevate whole blood NAD+ within 24 hours. However, there was an average increase in NAD+ with IV NAD+ at 7 and 14 days, though neither were statistically significant when compared to baseline levels or other treatment groups. It is hypothesized that the infusion of NAD+ results in a rise in extracellular NAD+, triggering an immune response that results in the adverse physical experiences (Adriouch *et al*., 2001; Liu *et al*., 2001). Though it is not clear why there would then be an increase in whole blood NAD+ at 7 and 14 days, such results were only mildly trending, and were not statistically significant compared to baseline or between group levels. For all groups, there was a mean whole blood NAD+ increase, which could be explained by an increase in hydration from the saline, for all but the oral group. The mechanisms resulting in the mild tingling experienced by the NR IV participants are not yet known, though the level of tingling appeared to be greater when the infusion rates were faster. NRIV resulted in a statistically significant increase in whole blood NAD+ compared to NAD+ IV and the saline control at 3 hours, and a statistically significant increase at 6 hours compared to saline, which was near significant (p=0.055) when comparing to NAD+ IV. The pharmacokinetics of NR IV and oral NR were different in this study, as IV NAD+ appeared to reach a maximum concentration (Cmax) at 3 hours, whereas oral NR Cmax was observed at the 24-hour timepoint.

This appears different than previous assessments, where a single oral dose of 1000 mg of NR resulted in a Cmax at 9 hours (n=1) in human peripheral blood mononuclear cells (Trammell *et al*. 2016a) and at 3 hours on Day 9 of a supplementation study (Airhart *et al*. 2017). These differences are likely explained by variations in dosing, biofluids sampled, and in the later study, protocol differences, as whole blood NAD was not measured on Day 1 in the naïve participants. Future clinical research will benefit from more study participants to improve the statistical power for these analyses.

## Limitations

An *a priori* statistical power analysis was not conducted for the current study, as this was a phase 0/1 assessment, and such head-to-head comparisons were not available in the literature in clinical or preclinical models. Though the population selected was determined to be healthy, a couple of participants were anemic/borderline anemic, and a couple had baseline elevated liver enzymes.

In clinical trials, controlling for hydration status when collecting serum samples is important, as most tests are a unit of measure, often weight or number per dilution status. (e.g., ng/dl or a number of cells per microliter). This control ensures the accuracy and reliability of the results. Additionally, the allowance of the consumption of food and beverages, while controlled, before the three-hour assessments confounded the results for glucose and insulin. The study results suggest that the NAD+ infusion may have resulted in an inflammatory response due to the increase in neutrophils 3 hours after the infusion in these participants. To validate these results, it is suggested that future studies incorporate additional clinical parameters of inflammation, including c-reactive protein, erythrocyte sedimentation rate, procalcitonin, calprotectin, and plasma viscosity, or the assessment inflammatory cytokines in plasma or serum.

Concerning the treatment dose, this study only compared a single infusion of each of the interventions, though in clinics, NAD+ may be offered at various doses from 250-1250 mg/day, as well as infused or injected over multiple days. To improve ecological validity, future studies would benefit from assessing multiple infusions as well as varying doses. In this study, mass equivalents of NR and NAD+ were utilized. The scientific community has not validated the presence of a cellular NAD+ transporter, thus NAD+ requires the release of its two phosphate groups, ultimately entering the cell as nicotinamide or nicotinamide riboside (Nikiforov *et al*., 2011). Direct comparison of molecular equivalents of NR and NAD+ is needed, as well as analyses of NAD+ from muscle and skin biopsies and other biofluids to determine why NAD+ IV failed to alter whole blood NAD+. It may also be beneficial to evaluate infusions followed by oral administration of NR to determine if oral supplementation is able to sustain elevations in NAD+ between infusion.

The NAD+ dried blood spots, while convenient for at home sampling, did not result in 100% usable samples. There were some initial challenges in understanding the instructions and ensuring that multiple blood drops did not touch in the designated locations on the cards. When blood samples overlapped or there were other deficits in the sample collection, these samples were unusable, and thus were excluded from the analysis.

## Conclusion

This is the first study to clinically evaluate nicotinamide riboside administered through an intravenous infusion. The acute intravenous infusions of 500 mg of pharmaceutically prepared NR IV were safe in the study participants, with minor infusion-related temporary experiences and no attributable adverse events for up to 14 days post-infusion. In comparison to NAD+ IV, NR IV was infused faster, and the infusion experience was more tolerable. NR IV increased NAD+ levels within 24 hours, which was surprisingly not observed with NAD+ IV. For future studies, is it recommended that protocols that provide multiple infusions of NAD+ are substituted with NR IV to determine if nicotinamide riboside can provide the same or greater benefits as NAD+ IV, but with fewer side effects and faster infusion rates.

## Data Availability

All data produced in the present study may be made available upon reasonable request sent to.

## Statement Contribution

- JH served as the contracted PI for this study, was responsible for IRB approval, final study protocol, supervising study conduct, data collection, data analysis, manuscript writing, editing, and final review of the manuscript.
- RI analyzed results, conducted statistical analysis, figure and table generation, and manuscript writing, editing, and final review of the manuscript.
- JK contributed to manuscript writing, data analysis, editing, and final review of the manuscript.
- AS was involved in the development of the study protocol, supervising study conduct, data collection, and manuscript revisions.
- ED was involved in study conduct, data collection, data interpretation, and review of the final manuscript.
- KD & EH contributed to study conduct, raw data outputs, data analysis, and final review of the manuscript.
- YNE was involved in the development of the study protocol, project management, study conduct, data collection, manuscript writing, editing, and final review of the manuscript.

### Acknowledgements

A special and huge thank you to Jaimee Arroyo and the staff and trainees at Hopewell Family Care Integrative Medicine and Roots Collaborative Care for generously providing the space and staff for making this study possible. Additionally, we would like to thank the amazing study participants; Dr. Ben Myatt at DCA Pharmacy, Nick Mitchell for getting us to the starting block; Dr. Chris Meletis for supporting data interpretation; and Albert Rembert for supporting the pre-study site assessment. We would also like to thank the team at Revvity for sample analysis of the dried blood spots.

## Abbreviations

CBC: complete blood count (hematology)
Cmax: maximum concentration
CMP: comprehensive metabolic panel (clinical chemistry)
DBP: diastolic blood pressure
DBS: dried blood spot
eNAD+: extracellular nicotinamide adenine dinucleotide
GRAS: generally recognized as safe
IV: intravenous
NAD+: nicotinamide adenine dinucleotide
NAD+ IV: intravenous NAD+
NADome: nicotinamide adenine dinucleotide precursors, intermediates, and metabolites
NMN: nicotinamide mononucleotide
NR: nicotinamide riboside
NR IV: intravenous NR
PARP: poly (ADP-ribose) polymerase
SBP: systolic blood pressure

## SUPPLEMENTAL TABLES

**Supplement Table 1.**
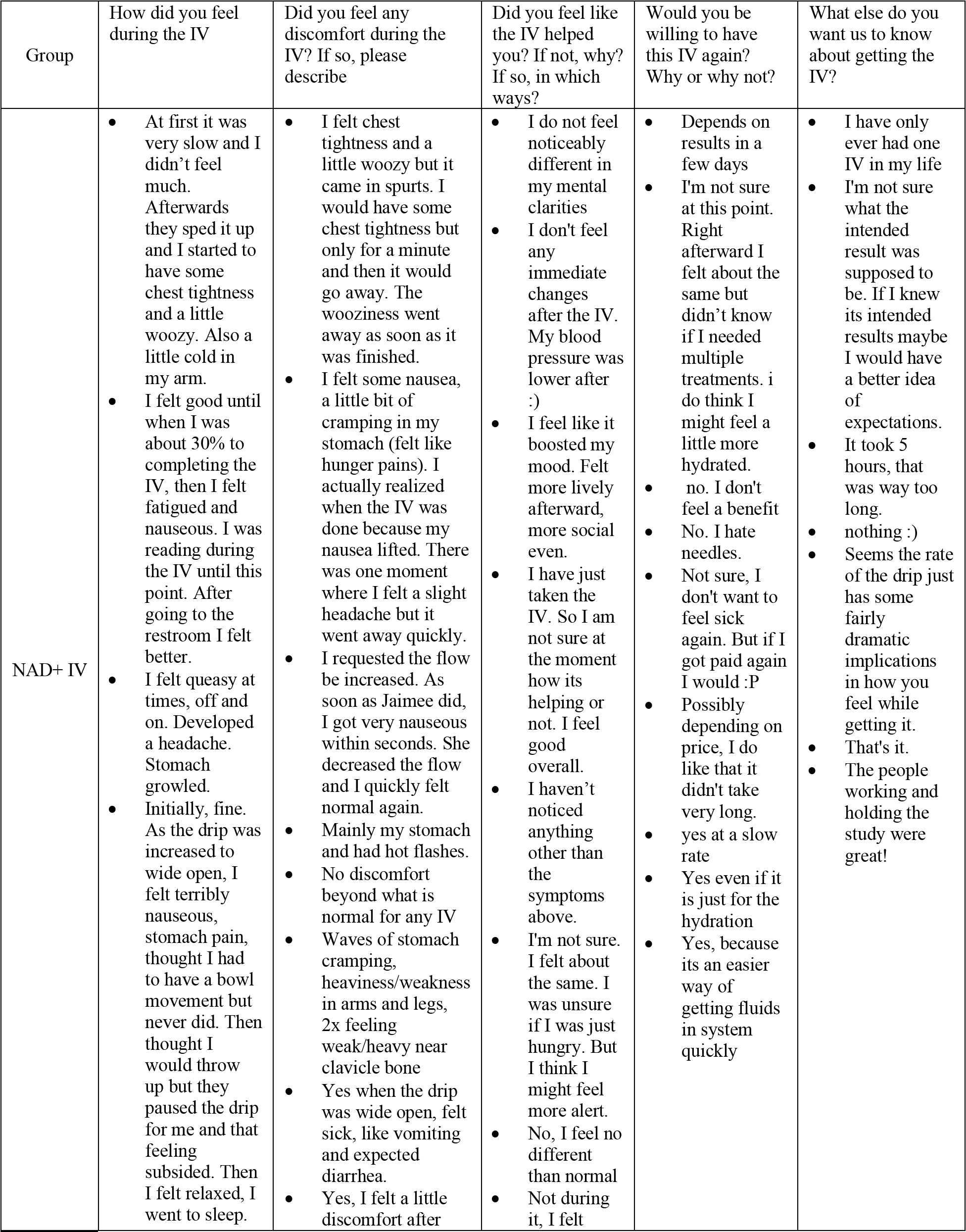
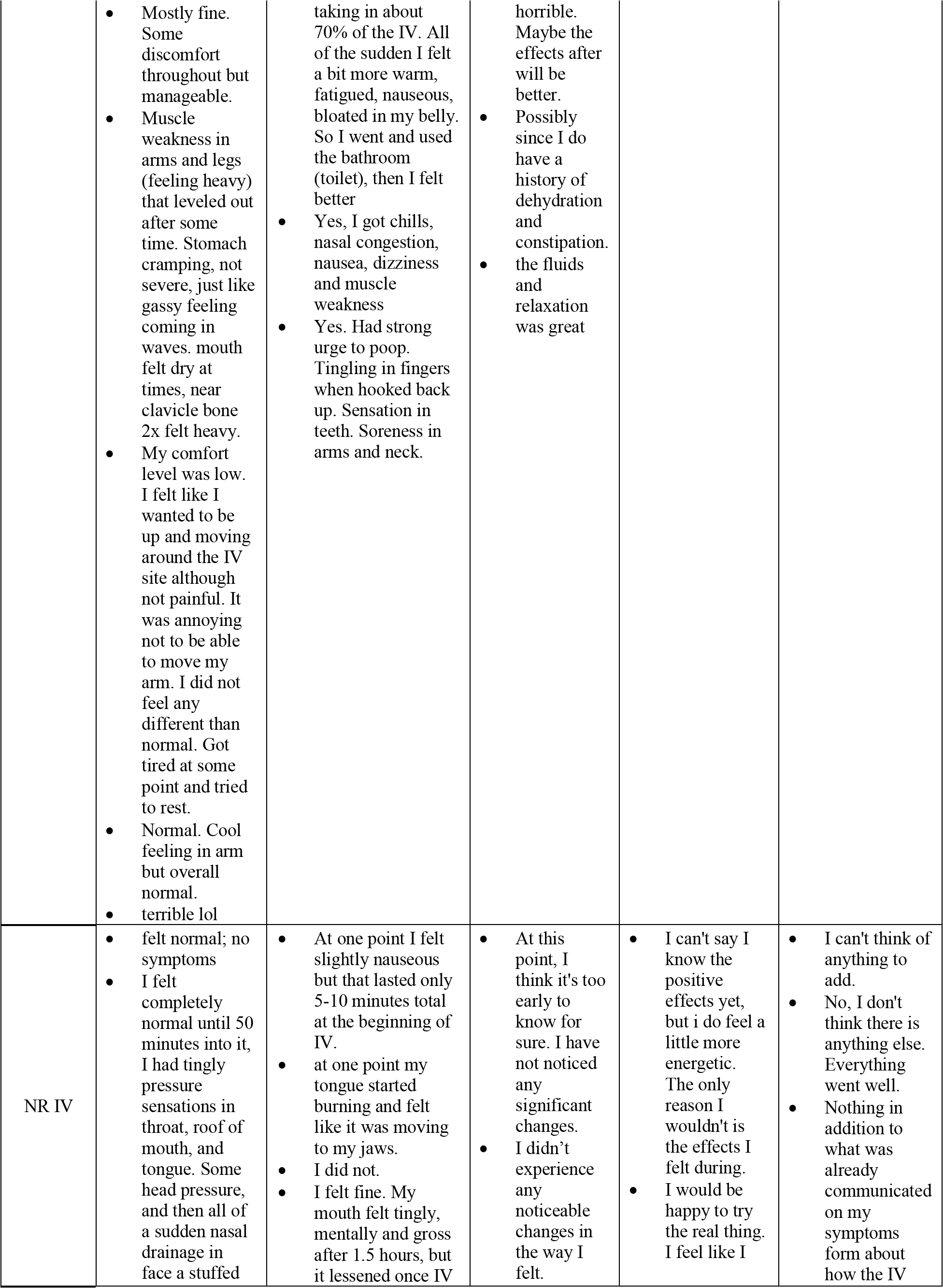
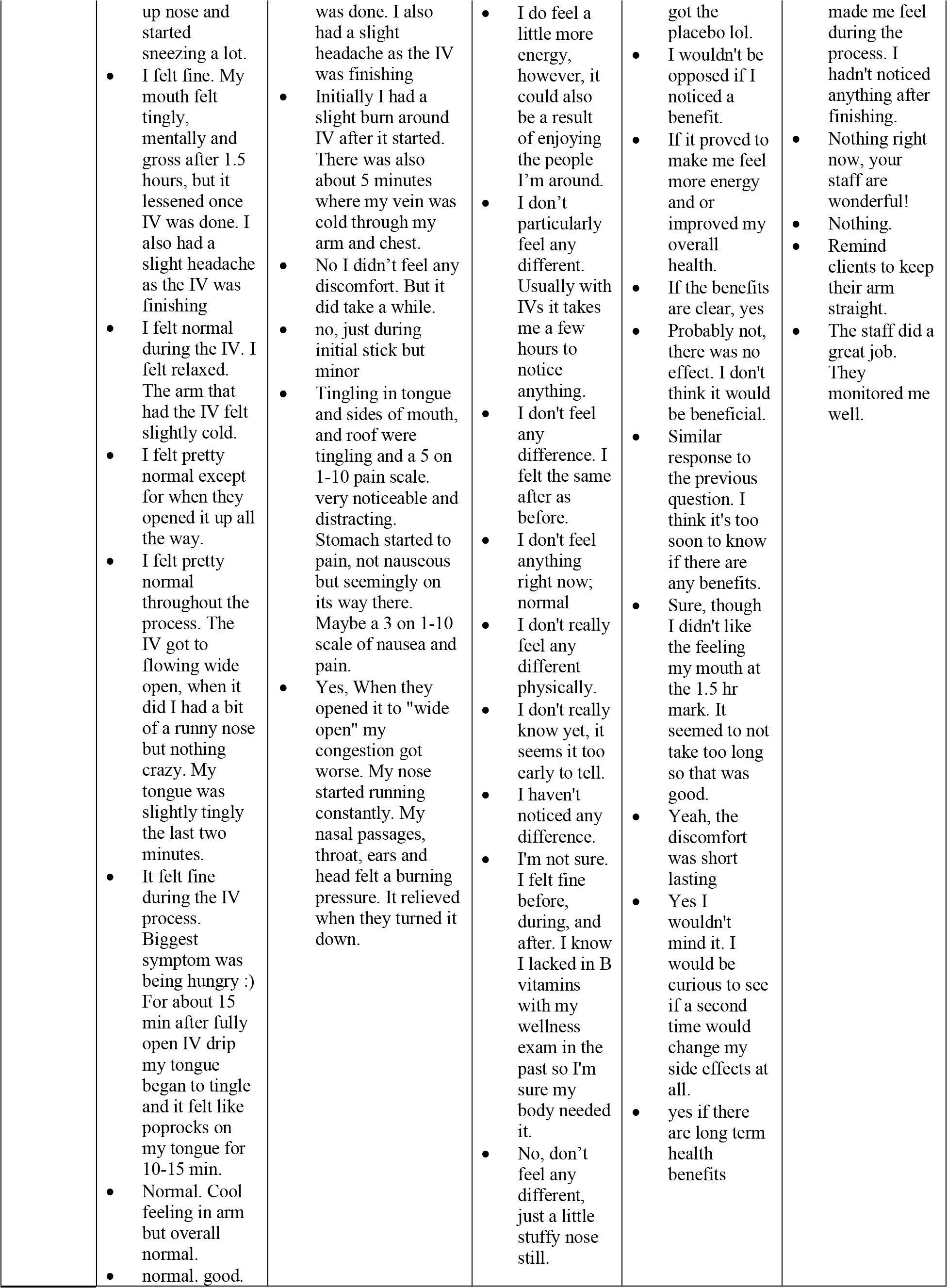
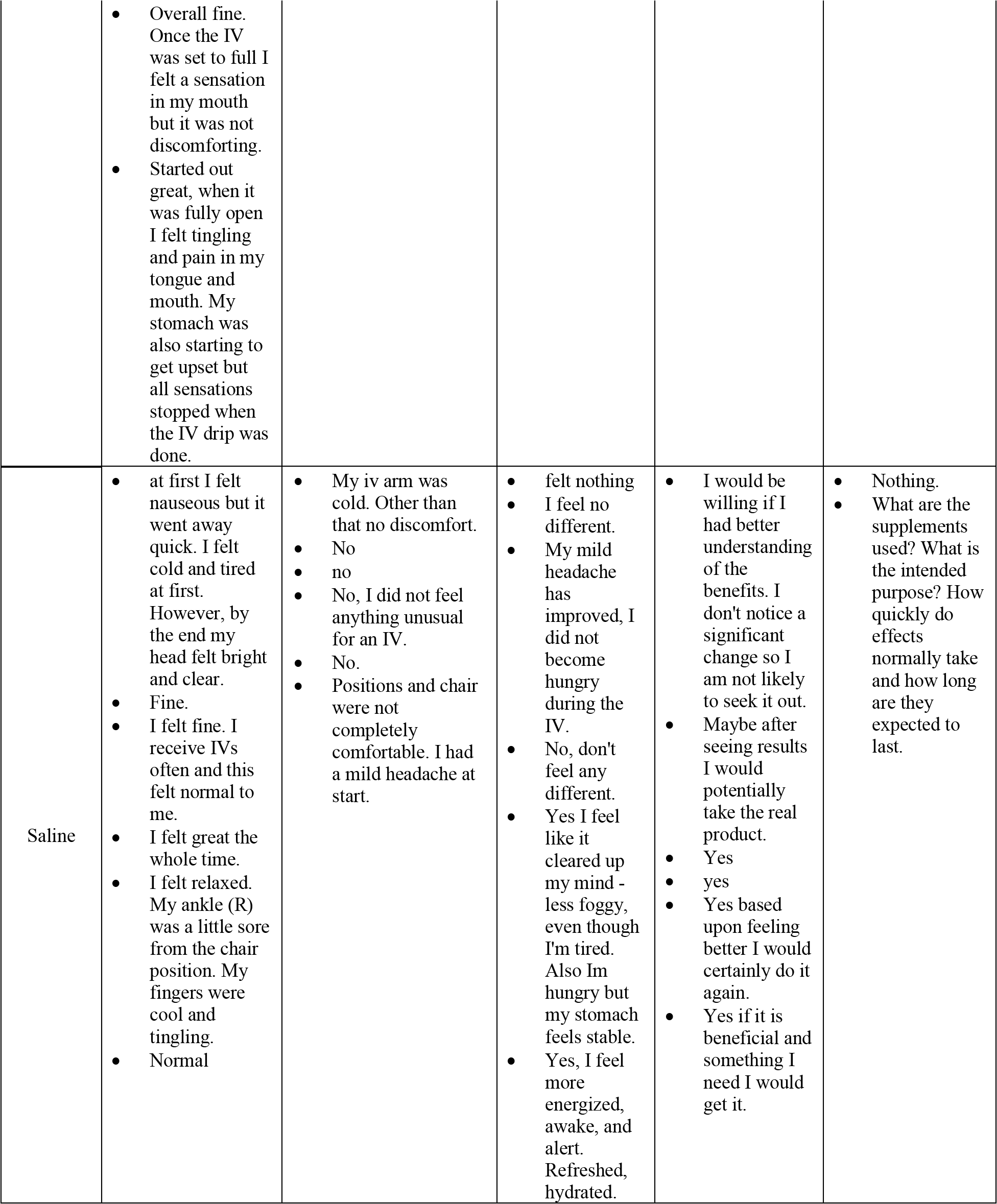
Study 1 Participant Described Experience.

**Supplement Table 2.** Study 2 participant experienced as documented by participants and monitoring health care providers.

| Group | Participant Experience | HCP Notes |
| --- | --- | --- |
| NAD+ IV | Aching all over, shortness of breath, chest pain; nausea, That was horrible. Haven't felt that bad since covid 2020 till she backed it off. Took 5-10 min to feel better, like how you feel right after throwing up euphoric, she turned it back up slightly- stomach pressure, nausea, shortness of breath.<br>Anxiousness; laid down; coming through, feeling gas in my stomach; neck and back ache wondering if I should have done this; i shouldn't have eaten such a big lunch; stomach kind of hurts; nurse slowed IV, eyes feel heavy; feels a little better since slowing the IV; feel better- muscle cramps between shoulder blades and neck has stopped mostly<br>Slight headache, bloating, flushed at first | Gets flushed above 125cc and it has to be turned down.<br>Had to have the IV taken all the way out because of discomfort and ran to the bathroom. Face was flushed.<br>Thought he was having a heart attack |
| NR IV | A little pressure/discomfort in jaws and sinus, faded within 10 min; "less groggy then when I arrived" tingling/discomfort in abdomen, faded after 5 min or so after finishing the IV.<br>Chill in the arm<br>Lip tingling<br>Stomach turned over, itchy throat and cough, tingling in right hand and fingers | Noted that he worked all day and said he was exhausted but is happy and surprised to find that he feels more clear headed and energized already |

